# Automated Phonological Error Scoring for Children with Language and Hearing Impairment

**DOI:** 10.1101/2024.09.04.24313011

**Authors:** Simon Sundström, Charalambos Themistocleous

## Abstract

**Purpose:** Phonological production impairments are prevalent in children with developmental language disorder (DLD) and hearing impairment (HI). This study aims to quantify and compare phonological errors in Swedish-speaking children using a novel automated assessment tool and provide an automatic machine learning classification algorithm of children with DLD and HI to age-matched controls based on phonological errors.

**Methods:** 72 Swedish-speaking children (29 with DLD, 14 with HI, and 29 typically developing) participated. Phonological production was elicited using a 72-item confrontation naming task. A novel tool was developed to calculate a composite phonological error score and specific scores for different phonological errors (deletions, insertions, substitutions, and transpositions) from written speech productions. This tool leverages the International Phonetic Alphabet (IPA) and a form of the Normalized Damerau–Levenshtein Distance for accurate error analysis.

**Results:** The composite score successfully differentiated between children with DLD and typically developing children, highlighting its sensitivity in detecting phonological impairment. Machine learning models can accurately differentiate between children with and without language disorders. However, children with DLD and HI differed in the phonemic deletion errors, which suggests that their phonemic production is relatively similar.

**Conclusions:** Children with DLD and HI exhibit significantly higher phonological error rates compared to typically developing peers. Children with HI and DLC are comparably impaired in phonology (as manifested by the compositive phonological score). These findings highlight the potential of machine learning for early identification and targeted intervention in language disorders, improving outcomes for affected children and demonstrated the potential of a multilingual tool for scoring phonological errors.

## 1 Introduction

Children with Developmental Language Disorder (DLD) experience difficulties in language production and perception, often appearing early in childhood and potentially persisting into adulthood. While the exact cause remains unknown, these challenges are primarily linked to neurocognitive factors. Similarly, children with hearing impairment (HI) often encounter difficulties in speech and language development. However, their challenges are directly related to sensory input limitations, referred to as language disorder associated with hearing loss (Bishop et al., 2017). Nevertheless, assuming the same prevalence of DLD as in the general pediatric population (at least 7%; Norbury et al. (2016)), a substantial proportion of children with HI can also be assumed to have language problems that are independent of their hearing ability, i.e., they have DLD, or would have had if not for their HI. This means that assessment procedures used with children with HI should be sensitive to whether language problems are related to their hearing impairment or to other factors (Hardman et al., 2023).

Both children with DLD (Lancaster & Camarata, 2019) and children with HI (Lederberg et al., 2013) demonstrate a wide range of individual differences in the severity and particular areas of language and communication affected. DLD can manifest as difficulties in expressive language (producing language) and/or receptive language (understanding language), impacting one or more linguistic domains, such as phonology (speech sounds), grammar (sentence structure), semantics (word meaning), or pragmatics (social use of language) (Bishop, 2017). Research has indicated that a substantial proportion (up to 40%) of children with DLD experience speech sound difficulties severe enough to warrant a diagnosis of speech sound disorder, with the majority (90%) of these cases being phonological in nature (Ttofari Eecen et al., 2019).

While many children with HI develop spoken language on par with typically developing peers, any degree of hearing loss poses a risk for delayed or atypical language development (Tomblin et al., 2015), especially in the domains of phonology and grammar, where impaired performance has been found in around 50% of children (Tuller & Delage, 2014). For example, both children with DLD and HI may experience difficulties in language perception, impacting their understanding of conversations and instructions. These language challenges can have a cascading effect on a child’s overall language abilities, impacting their verbal expression, vocabulary acquisition, and narrative skills. Frustration arising from communication difficulties can also lead to behavioral challenges in both DLD and HI populations. Not only does insufficient assessment and management of language difficulties in childhood and adolescence have consequences for communication but they likely also contribute to a higher prevalence of psychosocial difficulties and lower academic outcomes in both children with DLD (Botting et al., 2016; Ziegenfusz et al., 2022) and HI (de Jong et al., 2023; Sarant et al., 2015), hindering their education and impeding their academic success.

With respect to Swedish, both children with DLD and children with HI have been found to perform below their typically hearing peers on measures of phonological production accuracy. In a cross-sectional study of Swedish-speaking four–six-year-old children, Sundstrom et al. (2019) showed that children with DLD produced phonemes and stress patterns significantly less accurately compared to controls with typical language development. This was found for immediate repetition of real words and non-words, as well as for confrontation naming of pictures. Interestingly, no significant correlation was found between the repetition of stress and tonal word accents and measures like phonological production or receptive vocabulary Sundstrom et al. (2019). However, a significant correlation emerged between the ability to repeat stress in real words and expressive grammar skills.

Regarding Swedish-speaking children with HI, Nakeva Von Mentzer et al. (2015)investigated non-word repetition ability in five–seven-year-olds with severe bilateral HI who wore cochlear implants. Their findings suggested a lower ability of children with cochlear implants to repeat the consonants, vowels, primary stress and number of syllables in nonwords, compared to age-matched controls. Similar results for non-word repetition performance were obtained by Sundström et al. (2018b) in a study comparing four- to six-year-old children with moderate to severe HI who used cochlear implants or hearing aids to typically hearing age peers. In addition, they found that consonants, vowels and stress patterns were significantly harder for the children with HI to produce in a picture naming task (Sundström et al., 2018b).

Few studies have compared the phonological production ability of Swedish-speaking children with DLD and children with HI. Ibertsson et al. (2008) investigated nonword repetition ability in children aged 5–9 years. Consonant production accuracy were not found to differ between children with mild–moderate HI who used hearing aids and children with SLI, while children with cochlear implants performed significantly below the other groups. In a more recent study, Sundström et al. (2018a) found no difference between children with DLD and children with HI in the ability to correctly produce consonants and vowels when repeating words and nonwords, or when naming pictures.

In summary, previous studies indicate that Swedish-speaking children with DLD and children with HI may face considerable challenges in accurately producing consonants, vowels, stress and tonal word accents. However, differentiating between these groups based on phonological production performance may require more in-depth error analyses, rather than coarse judgments of speech sounds as either correct or incorrect.

Consequently, screening and assessment procedures for children with HI need to be capable of differentiating between language difficulties stemming from their hearing impairment and those arising from other underlying factors, such as DLD (Hardman et al., 2023). This distinction is crucial because it has implications for both diagnosis and treatment. If a child’s language difficulties are primarily attributable to their hearing impairment, interventions focused on improving auditory access and providing appropriate language models will be most beneficial. However, if DLD is also present, additional therapeutic approaches targeting specific language skills and cognitive processes may be necessary. Furthermore, recognizing the potential coexistence of DLD and HI highlights the importance of comprehensive assessment practices for children with hearing loss. By employing tools and techniques sensitive to both auditory and language processing, clinicians can gain a more accurate understanding of a child’s unique needs and develop tailored intervention plans that address their specific challenges, ultimately promoting optimal language development outcomes. The present study focuses on an aspect of language use that is often problematic for children with DLD and children with HI, namely phonological production, defined as the ability to correctly produce speech sounds. Studying phonological errors in these groups may yield important insights into not only what characterizes phonological production ability in each group, but also concerning similarities and differences.

### The present study

The manual scoring of phonological errors presents significant challenges within clinical and research settings. It is a labor-intensive and time-consuming process, often susceptible to human error and variability. Accurate and reliable analyses are contingent upon clinicians possessing specialized expertise in phonetic transcription and a nuanced understanding of the speaker’s dialectal variations (Kent, 1996). These demands can strain resources and potentially introduce inconsistencies in assessment outcomes. To address these limitations earlier research has employed distance metrics to provide an Error Frequency Analysis in individuals with acquired language disorders, such as apraxia of speech and aphasia with phonemic paraphasia (Smith et al., 2019) and to score spelling differences in individuals with PPA (Themistocleous et al., 2020). For example, in our previous study, Themistocleous et al. (2020) compared and evaluated several distance metrics and showed that the *Normalized Damerau–Levenshtein Distance* replicates the manual scoring process. The advantage of the *Normalized Damerau–Levenshtein Distance* over other distance metrics is that it accounts for the transpositions of phonemes or graphemes, whereas the simpler *Levenshtein Distance* only for deletion, insertion, and substitution errors.

To enhance the efficiency and objectivity of phonological assessment, this study proposes an automated Composite Phonological Score (CPS) specifically designed for evaluating the phonological performance of children with DLD. The CPS algorithm leverages a string distance metric, the Normalized Damerau–Levenshtein Distance (Jurafsky & Martin, 2009), to compute the phonemic dissimilarities between a child’s production and the target utterance. This approach enables the quantification of phonological accuracy at a phonemic level. We have refined the algorithm to generate not only a composite score but also discrete scores for specific categories of phonetic errors, namely deletions, insertions, substitutions, and transpositions. This metric compares the response word and the target word. To account for phonemic processes that appear at the surface level, we transcribe these both words in the International Phonetic Alphabet (IPA). The IPA transcription involves both the representation of phonemes into words and phonological processes involved in the production of the surface phonemic forms. In Swedish, for example, the velar /k/ undergoes a change to become alveopalatal /ɕ/ when it appears before /i/ or /y/, /ø/, /ä/. This can be observed in words like *kille* (/ˈɕɪlːɛ/) or *köpa* (/ˈɕøːpa/). The phonemic distance is calculated using the *Normalized Damerau–Levenshtein Distance,* which accounts for four types of phonemic errors, namely, deletions, insertions, substitutions, and transpositions of phonemes (Themistocleous et al., 2020). A multilingual web tool, utilizing the *Phonemic Distance Algorithm*, offers support for scoring phonological errors in over 65 languages and language varieties, and is available at Open Brain AI (http://openbrainai.com) (Themistocleous, 2024). The final score is a distance metric from 0 when the words are identical to 1 when they are dissimilar. This scoring system provides clinicians and researchers with a more comprehensive understanding of a child’s phonological profile, facilitating targeted intervention planning and progress monitoring.

This study has two primary objectives. First, we aim to quantify phonological production differences in Swedish-speaking children with DLD and HI compared to Typically Developing (TD) children. By establishing measurable benchmarks, we hope to improve early diagnosis and intervention for these conditions. Second, we intend to develop a classification model to differentiate between children with DLD, HI, and aged matched typically developing children based on their phonological production patterns. Our primary hypothesis is that Swedish-speaking children with DLD and HI will exhibit distinct phonological production profiles compared to TD children and differ from each other. To evaluate this, we have designed a two-step machine learning approach (Figure 1):

(1) *Classify TD vs. non-TD children:* This initial step differentiates typically developing children from those with either DLD or HI.
(2) *Classify DLD vs. HI:* The second step focuses on distinguishing between children with DLD and HI.

**Figure 1.**
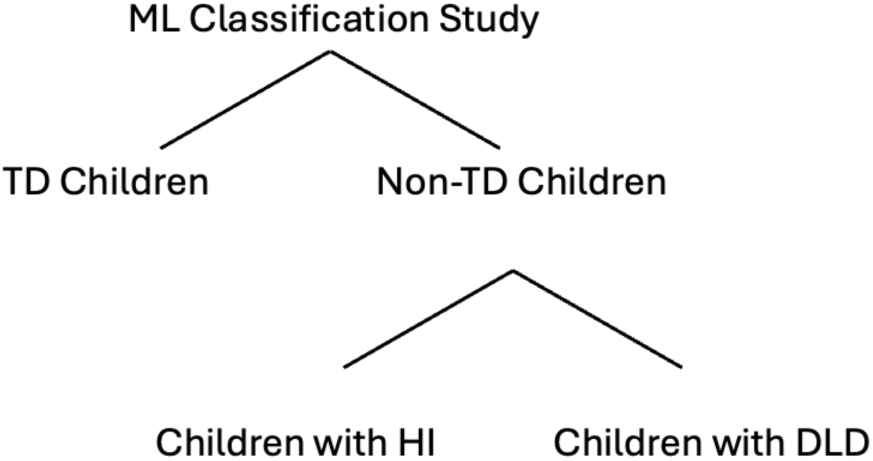
Two-fold ML Classification Approach: (1) distinguishing between typically developing children (TD) and non-TD children; subsequently (2) classifying between children with HI and children with DLS.

If successful, this automated system has the potential to expedite diagnosis, allowing clinicians to quickly identify and address phonological difficulties, which is particularly vital for early and effective intervention in language impairment cases.

## 2 Methodology

### 2.1 Participants

72 children, 29 with DLD, 14 children with HI and 29 age-matched, typically developing children, participated in the study. All children were between 4 and 6 years old, speakers of the Central Swedish dialect, and had no known neurodevelopmental or psychiatric disorders. Children with DLD received a DLD diagnosis by a speech and language therapist and had mainly phonological and grammatical, excluding pronounced lexical, semantic, and pragmatic deficits. The children with HI all had congenital or early acquired bilateral mild to profound sensorineural hearing loss.

Three of the children had bilateral cochlear implants, nine used bilateral conventional air conduction hearing aids, and two had a bimodal solution with a hearing aid in one ear and a cochlear implant in the other.

#### 2.1.1 Age in Months

Given the rapid cognitive and linguistic changes in early life stages such as infancy and early childhood, sage in months allows for a more nuanced understanding of development.

#### 2.1.2 Maternal Education Years

Maternal Education Years is a proxy for the home educational environment or socioeconomic status. This metric aims to control for environmental variables that could influence cognitive or linguistic development.

#### 2.1.3 Non-Verbal Cognitive Ability

The Cognitive Ability score is a metric of non-verbal intelligence that attempts to encapsulate an individual’s general cognitive functions. It is assessed through the block design subtest of the Swedish version of the Wechsler Preschool and Primary Scale of Intelligence, Third Edition (WPPSI–III) (Wechsler, 2005) a standardized intelligence test.

#### 2.1.4 Word and Nonword Repetition

Word and Nonword Repetition evaluates phonological memory and language processing abilities. The individual is tasked with repeating real or made-up words, and the proportion of accurately repeated phonemes is calculated. This measure is often used to diagnose children with DLD and other language disorders.

#### 2.1.5 Expressive Grammar

Expressive Grammar was assessed with Gramba (Hansson & Nettelbladt, 2004). The ability to use noun morphology, verb morphology, and syntax are probed through a sentence completion test. During this assessment, children are prompted to complete sentences, allowing researchers to gauge their grasp of these fundamental aspects of language. To ensure accuracy and enable detailed analysis, all responses were audio-recorded and later transcribed. The transcriptions were then scored, with a maximum achievable score of 44. This scoring system provides a quantifiable measure that enables the comparison of linguistic abilities across individuals or groups.

#### 2.1.6 Receptive Vocabulary Raw

Receptive Vocabulary Raw scores aim to quantify the breadth of words an individual can understand. Receptive Vocabulary was assessed using The Peabody Picture Vocabulary Test, or PPVT–III, adapted in Swedish. The examiner pronounces a word, and the child is asked to point to one of four pictures that best represents the meaning of that spoken word. Given the absence of standardized norms for Swedish children between the ages of four and six, the study relies on the raw max scores for analysis.

### 2.2 Procedures and Phonological Scoring

#### 2.2.1 72-item confrontation naming task

Children were assessed for language impairment during one or two sessions (total duration 90– 120 minutes including suitable breaks). The testing took place in a quiet room at Linköping University hospital, at the preschool, or in the child’s home. All testing was administered by the first author. Phonological production was elicited with a 72-item confrontation naming task. In this task, researchers assessed participants’ ability to produce sounds and words accurately by presenting them with pictures of objects and asking them to name the object depicted (Sundstrom et al., 2019).

#### 2.2.2 Automated Phonological Scoring

The Normalized Damerau–Levenshtein Distance is a distance metric, which allows the comparison of two strings and calculates a composite score of the differences between the two strings. A string can be a sequence of letters (graphemes), phonemes or another type of elements. The Damerau–Levenshtein function (*dlev_a,b_*(*i*, *j*)) is the distance between *two strings of phonemes’ words’ a* and *b* by estimating the distance between the *i*–symbol of an initial substring of the word *a* (prefix) and a *j*–symbol prefix of word *b*. The *restricted distance* is calculated recursively as:

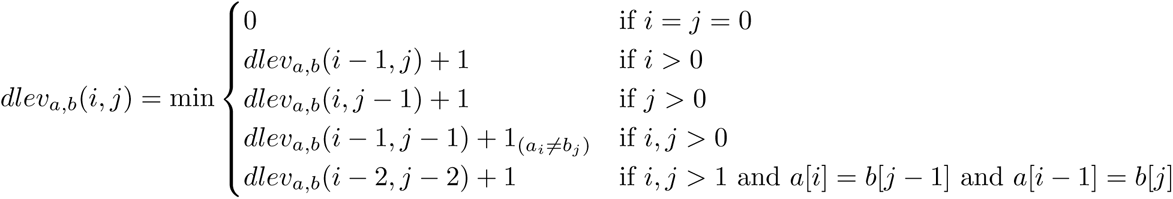

*where* 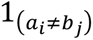 *is equal to 0 when a_i_* = *b_j_ and is equal to 1 otherwise. The phonemic scoring requires the match one of the following processes:*

1. *dlev_a,b_*(*i* − 1, *j*) + 1: a deletion from word a to word b.
2. *dlev_a,b_*(*i*, *j* − 1) + 1: an insertion from word a to word b
3. 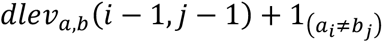: a (mis-)match from word a to word b.
4. *dlev_a,b_*(*i* − 2, *j* − 2) + 1: transposition

Typically, a score of 0 indicates perfect phonological production (i.e., no difference between the target and the response), and higher scores indicate more errors.

For the purposes of this study, we have adapted the Normalized Damerau–Levenshtein Distance algorithm to automatically compute the *Composite Phonological Score of Errors (CPS)* metric from written speech productions. This adaptation enables the identification and categorization of specific speech errors, *deletions* (i.e., the absence of phonemes that should be present in the target production), *insertions* (i.e., the presence of extra phonemes not found in the target production; the s*ubstitutions* (i.e., the replacement of one phoneme with another), and *transpositions* (i.e., the swapping of the order of two adjacent phonemes).

Our implementation involves a two-step process. The first step involves the Phonetic Transcription. The written speech productions are first converted into their corresponding representations using the *International Phonetic Alphabet* (IPA). This standardized phonetic notation allows for a precise comparison at the sound level, regardless of the original language or orthography. The second step involves the Error Calculation. The IPA transcription of the target production is then compared to the IPA transcription of the response. The modified Normalized Damerau–Levenshtein Distance algorithm analyzes the differences between these two phonetic sequences, identifying and classifying the specific error types (deletions, insertions, substitutions, and transpositions). This information is then used to calculate the CPS metric, providing a quantitative assessment of the accuracy of the speech production. The Automated Phonological Scoring is approach was developed and implemented in *Open Brain AI* (http://openbrainai.com) (Themistocleous, 2024) and provides a Multilingual Composite Phonological Score of Errors and the individual phonemic errors adapted to several languages.

#### 2.2.3 Statistical Analysis

Pearson and Spearman rank-order correlations were computed between CPS, phonemic errors, and the other numerical variables of interest. Statistical tests were conducted to assess the differences between the groups. The choice of statistical test (independent *t*-test or Mann-Whitney U) was determined based on the Levene’s test for equality of variances.

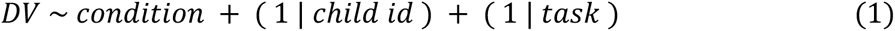

We employed linear mixed-effects models to evaluate the effects of children with DLD vs. typically developing children on the phonological distance. The dependent variable was the phonological distance and the *condition* (children with DLD vs. typically developing children) were the predictors. The *participant* was modeled as a random slope to model participants’ differences. We computed the Linear Mixed-Effects Models in R, a programming language for statistical analysis (R Core Team, 2020), using the “lme4” package (Bates et al., 2015) and extracted the p values with the LmerTest package (Kuznetsova et al., 2016). Finally, post-hoc contrasts were provided using the R package emmeans (EMMs, also known as least-squares means), which provides estimated marginal means (Russell, 2020). A *t*-test was performed to compare the duration of vowels and consonants produced by patients and clinicians.

### 2.3 Machine Learning

From our statistical models, we wanted to assess the implications of our measures for identifying a child with DLD from children with TD. Therefore, we designed a robust classification approach to work with a small amount of data. To assess the diagnostic value of phonological scores as predictors of hearing Impairment and DLD, we implemented two machine learning approaches. The first approach focused on differentiating typically developing children from those with hearing Impairment or DLD based on their phonological errors. Subsequently, we explored the ability to differentiate within the latter group, specifically between children with DLD and those with hearing Impairment.

#### 2.3.1 Model Comparison

For the classification we evaluated three classifiers: Decision Tree Classifier, Random Forest Classifier, and Gradient Boosting Classifier. Each of these classifiers has unique characteristics and is suited for several types of data and classification problems. The primary motivation for using these models is that they are able to support smaller datasets and also provide feature importance, which was critical for us because we wanted to assess the phonological measures developed for this study.

*1. Decision Tree Classifier (DT)* is a simple, yet powerful model used in supervised learning. For the classification it creates a tree-like graph of decisions by spliting the data into branches at decision nodes, which are formed based on feature values. Each decision node in the tree represents a feature in an instance to be classified, and each branch represents a value that the node can assume. Instances are classified starting from the root node and sorted based on their feature values down the tree along the branches until they reach a leaf node, which provides the classification (Hastie et al., 2009).
*2. Random Forest Classifier (RF)* this is an ensemble learning method for classification that operates by constructing a multitude of DTs at training time and outputting the class that is the mode of the classes of the individual trees (Breiman, 2001). In other words, RFs create a set of decision trees from randomly selected subsets of the training set, then aggregate their individual predictions to make a final decision. It introduces additional randomness when growing trees; instead of searching for the most important feature while splitting a node, it searches for the best feature among a random subset of features. Because of its ensemble nature, the random forest can achieve higher accuracy than a single DT and is better at avoiding overfitting. Random forests handle unbalanced and missing data and maintain accuracy for missing data.
*3. Gradient Boosting Classifier (GBC)* is an ensemble approach, like RFs, that builds models sequentially, each new model attempting to correct errors made by the previous ones (Bentéjac et al., 2021). Models are added one at a time, and existing models in the sequence are not changed. It involves three parts: a Impairment function to be optimized, a weak learner to make predictions, and an additive model to add weak learners to minimize the Impairment function. New learners are created to be maximally correlated with the negative gradient of the Impairment function associated with the whole ensemble.

#### 2.3.2 Preprocessing and Addressing Data Imbalance

For the ML classification, we employed the automated phonological error scores for each word (CPS and error types), demographic data (age and maternal education), and psychological assessments (nonverbal IQ, word and nonword repetition abilities, and vocabulary skills) (Table 1). The individual scores of each word items of the test were consolidated into a single score performance for each child by calculating median values for each child. Following data preparation, we addressed potential imbalances in how frequently each condition occurs within their dataset. Balancing a dataset is crucial for developing a reliable and fair predictive model as class imbalance can lead the machine learning model to develop a bias toward the majority class, often at the expense of poorly predicting the minority class. To counter this imbalance, we employed SMOTE and works by creating samples by over-sampling with replacement (Chawla et al., 2002). During *five*-fold cross-validation, SMOTE is applied within each training fold separately, ensuring that each fold used for validation remains a true representation of the original dataset. This approach provides a robust estimate of how the model will perform on unseen data. We set up a machine learning pipeline to manage missing data (using median values to fill in gaps), standardize measurements (to give them equal weight in the analysis), and apply the three classifications classification algorithms to predict the condition based on the input data.

**Table 1.**
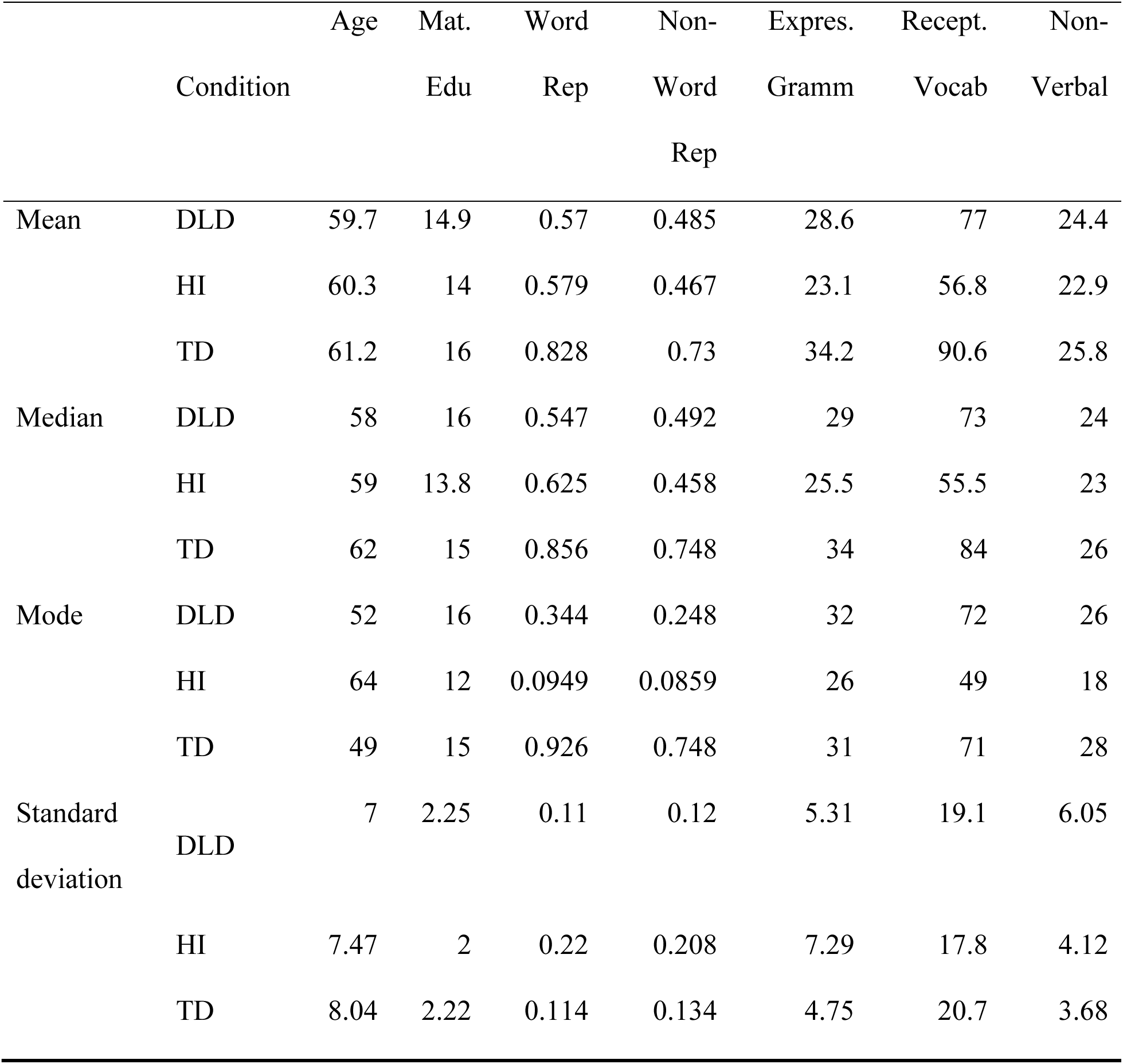
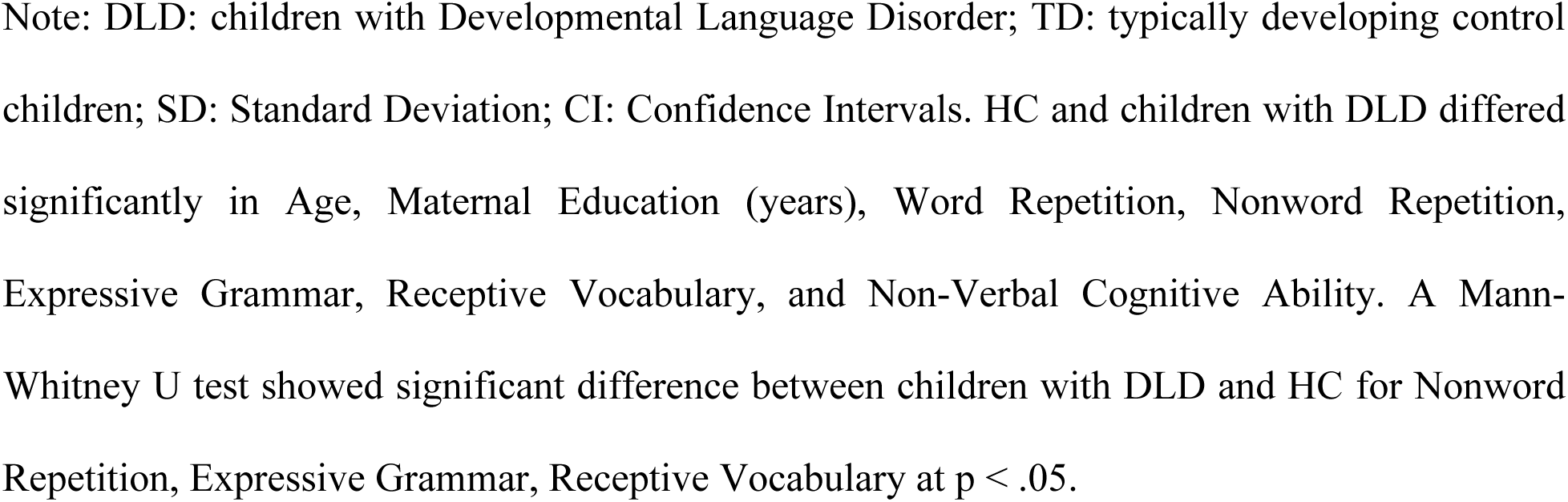
Demographic information of the participants for each group children with DLD, HI, and TD children for age, maternal education and their scores in neurolinguistic testing.

|  | Condition | Age | Mat. Edu | Word Rep | Non-Word Rep | Expres. Gramm | Recept. Vocab | Non-Verbal |
| --- | --- | --- | --- | --- | --- | --- | --- | --- |
| Mean | DLD | 59.7 | 14.9 | 0.57 | 0.485 | 28.6 | 77 | 24.4 |
|  | HI | 60.3 | 14 | 0.579 | 0.467 | 23.1 | 56.8 | 22.9 |
|  | TD | 61.2 | 16 | 0.828 | 0.73 | 34.2 | 90.6 | 25.8 |
| Median | DLD | 58 | 16 | 0.547 | 0.492 | 29 | 73 | 24 |
|  | HI | 59 | 13.8 | 0.625 | 0.458 | 25.5 | 55.5 | 23 |
|  | TD | 62 | 15 | 0.856 | 0.748 | 34 | 84 | 26 |
| Mode | DLD | 52 | 16 | 0.344 | 0.248 | 32 | 72 | 26 |
|  | HI | 64 | 12 | 0.0949 | 0.0859 | 26 | 49 | 18 |
|  | TD | 49 | 15 | 0.926 | 0.748 | 31 | 71 | 28 |
| Standard deviation | DLD | 7 | 2.25 | 0.11 | 0.12 | 5.31 | 19.1 | 6.05 |
|  | HI | 7.47 | 2 | 0.22 | 0.208 | 7.29 | 17.8 | 4.12 |
|  | TD | 8.04 | 2.22 | 0.114 | 0.134 | 4.75 | 20.7 | 3.68 |

Note: DLD: children with Developmental Language Disorder; TD: typically developing control children; SD: Standard Deviation; CI: Confidence Intervals. HC and children with DLD differed significantly in Age, Maternal Education (years), Word Repetition, Nonword Repetition, Expressive Grammar, Receptive Vocabulary, and Non-Verbal Cognitive Ability. A Mann-Whitney U test showed significant difference between children with DLD and HC for Nonword Repetition, Expressive Grammar, Receptive Vocabulary at $p < .05$ .

#### 2.3.3 Training the models

The training of the models was conducted using a five-fold grouped cross-validation, which considers the child id to ensure that there is no data leakage from the training test into the test set. To tune the models and select the best one employed GridSearchCV, an approach implemented in scikit-learn that automate the process of tuning parameters of a model by evaluating multiple combinations of parameter tunes, cross-validate the results to find the best set of parameters that offer the most accurate model.

A 5-fold cross-validation was used to split into 5 different subsets; each fold serves as a test set once, with the remaining parts used as the training set, thus rotating through all combinations. The performance of the final model is evaluated through standard statistical measures, namely their accuracy, F1 score, and by generating a confusion matrix and a classification report that explain how well the model predicts conditions and where it might be making errors. We then extracted the feature importance from the final ML model. Each classifier was trained using the training data from the group five-fold and then validated on the testing data. Post prediction, each model’s performance was assessed using standard evaluation metrics, namely the *accuracy*, which is the proportion of correctly predicted classification; the F1 Score, which is the harmonic mean of precision and recall; the precision, which is the proportion of positive identifications that were indeed correct; the recall, which is the proportion of actual positives that were identified correctly; the Area Under the Receiver Operating Characteristic Curve (ROC/AUC), and the Cohen’s Kappa (Hastie et al., 2009).

## 3 Results

In this section, first we discuss the differences between children with DLD, HI, and TD. Table 2 shows the Mean and Standard Deviation (SD) scores. Subsequently, we present the statistical analysis for each of the outcome phonological measures.

**Table 2.**
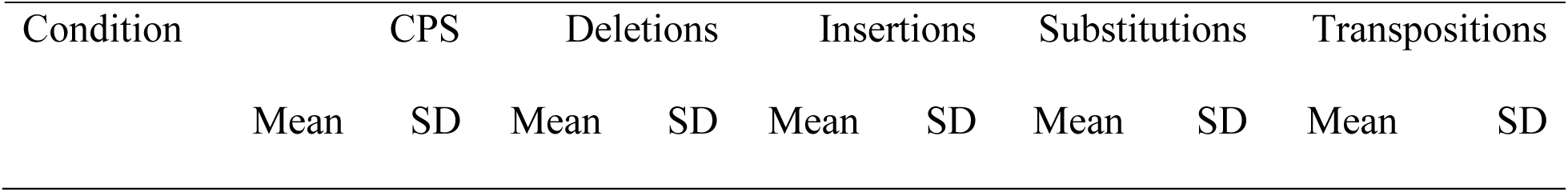
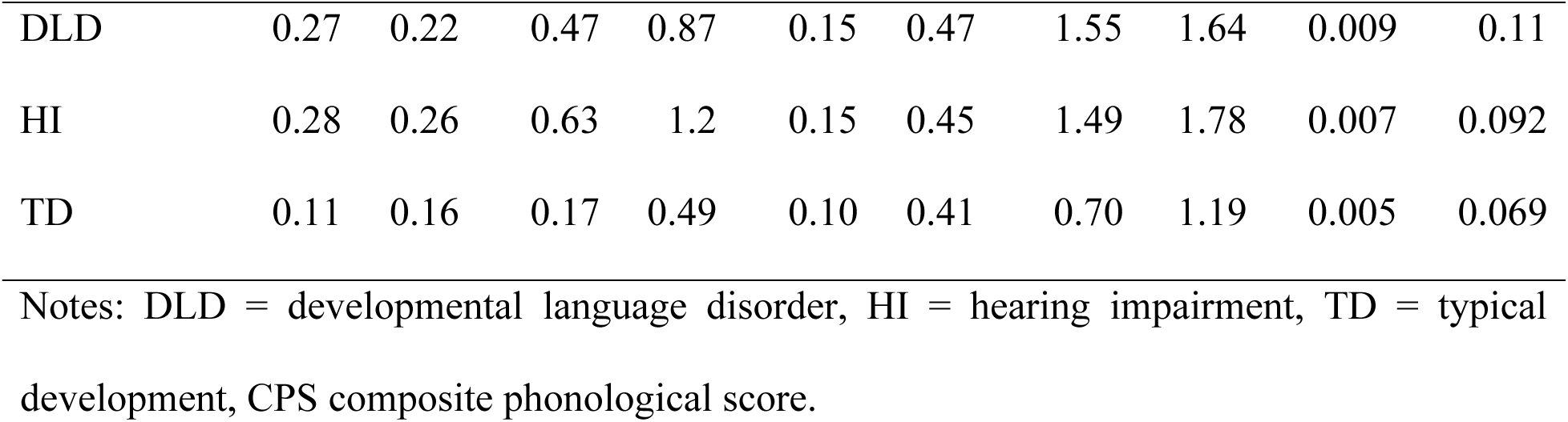
Mean and Standard Deviation (SD) scores.

Across all three groups (DLD, HI, and TD), substitutions are the most frequent type of phonemic error. Children with DLD and HI exhibit a similar rate of substitutions, which is notably higher than that of TD children. Deletions are the next most frequent error type, with the HI group showing the highest rate, followed by DLD, and then TD. Insertions occur at a relatively low rate across all groups, with DLD and HI children showing similar rates, slightly higher than TD children. Transpositions are the least common error, with minimal occurrences in all groups. Overall, children with DLD and HI demonstrate considerably higher rates of phonemic errors compared to TD children, suggesting greater challenges in phonological development.

### 3.1 Phonological Errors: CPS, Deletions, Insertions, Substitutions, Transpositions

Table 3 shows the comparative effects of the three groups, namely DLD, HI, and TD on the CPS.

**Table 3.**
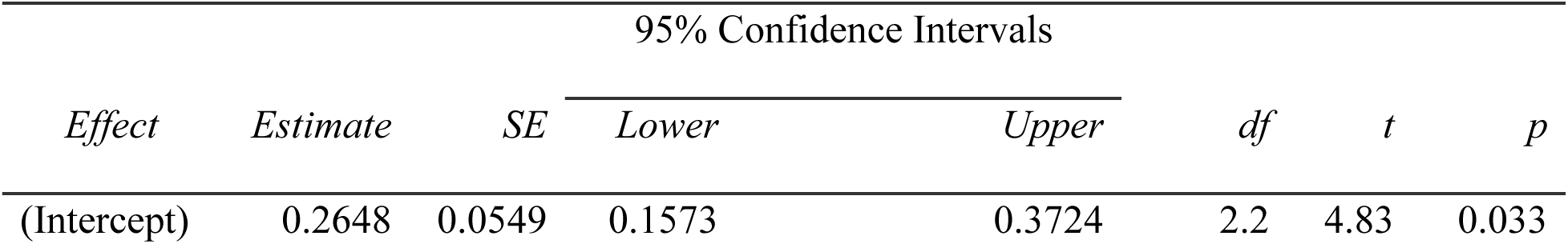
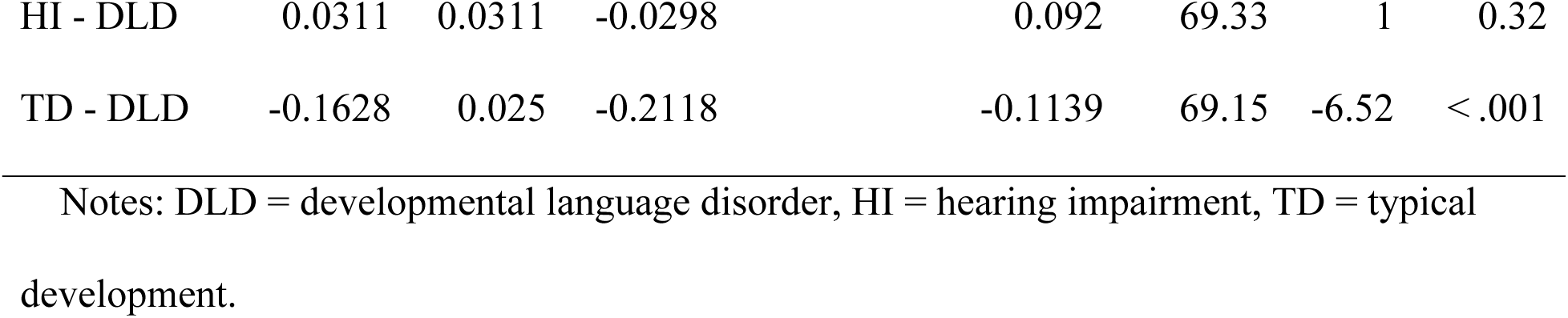
Statistical analysis of the effect of the group on the CPS.

| *Effect* | *Estimate* | *SE* | 95% Confidence Intervals |  | *df* | *t* | *p* |
| --- | --- | --- | --- | --- | --- | --- | --- |
|  |  |  | *Lower* | *Upper* |  |  |  |
| (Intercept) | 0.2648 | 0.0549 | 0.1573 | 0.3724 | 2.2 | 4.83 | 0.033 |
| HI - DLD | 0.0311 | 0.0311 | -0.0298 | 0.092 | 69.33 | 1 | 0.32 |
| TD - DLD | -0.1628 | 0.025 | -0.2118 | -0.1139 | 69.15 | -6.52 | < .001 |

Notes: DLD = developmental language disorder, HI = hearing impairment, TD = typical development.

TD compared to children with DLD and HI differed significantly in their composite phonological score. Children with TD produced phonological errors closer to 0 whereas both children with DLD and HI had a composite phonological score closer to 30%. The difference in the productions was also shown in the post-hoc Bonferroni analysis, which showed significant differences between children with DLD and TD (p < .001) and HI and TD (p < .001) but no difference between children with DLD and HI.

#### 3.1.1 Deletions

Table 4 shows the results of the statistical analysis of the effects of children with DLD, HI, and TD on the phonemic deletion errors.

**Table 4.**
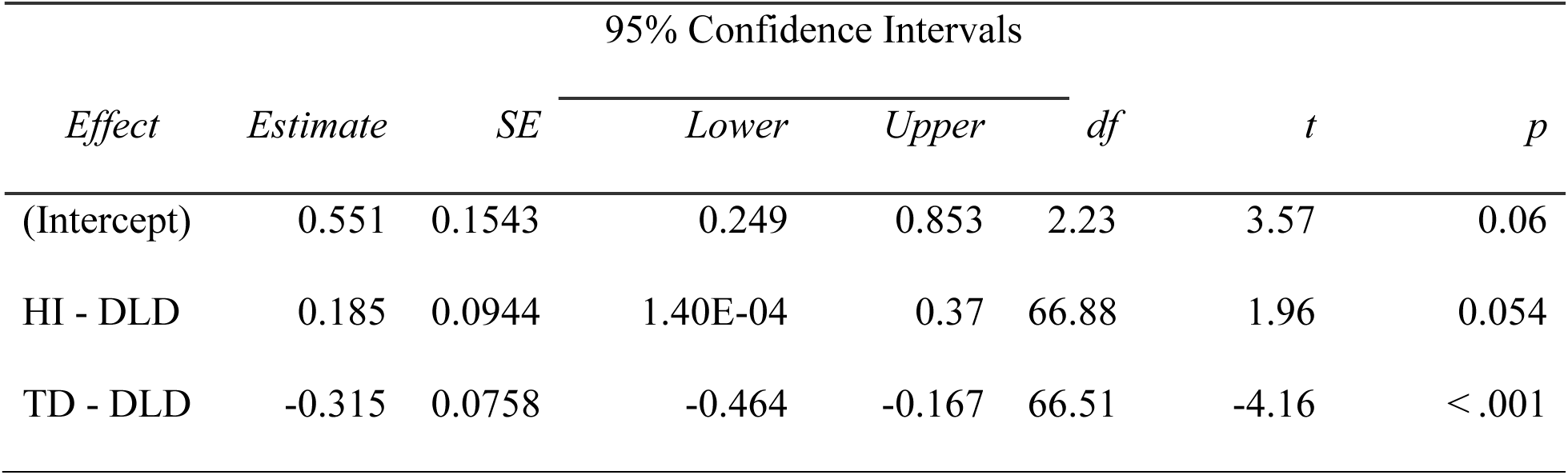
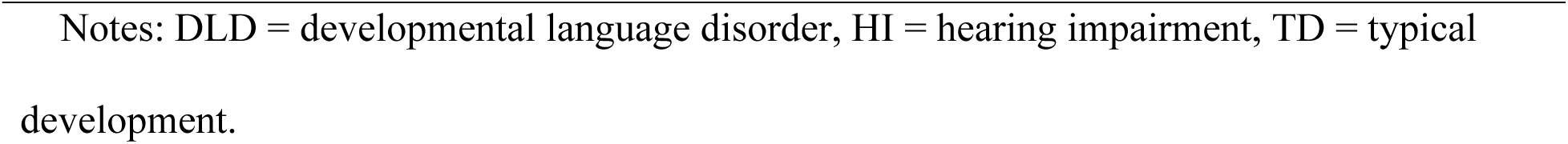
Statistical analysis of the effect of the group on the deletions.

| *Effect* | *Estimate* | *SE* | 95% Confidence Intervals |  | *df* | *t* | *p* |
| --- | --- | --- | --- | --- | --- | --- | --- |
|  |  |  | *Lower* | *Upper* |  |  |  |
| (Intercept) | 0.551 | 0.1543 | 0.249 | 0.853 | 2.23 | 3.57 | 0.06 |
| HI - DLD | 0.185 | 0.0944 | 1.40E-04 | 0.37 | 66.88 | 1.96 | 0.054 |
| TD - DLD | -0.315 | 0.0758 | -0.464 | -0.167 | 66.51 | -4.16 | < .001 |

Notes: DLD = developmental language disorder, HI = hearing impairment, TD = typical development.

Children with HI and DLD differed significantly from the TD children regarding deletions, which is also evidenced by the higher number of deletions in children with DLD and HI compared to TD as shown in Table 4. Specifically, we find that deletions occur more frequently in children with HI compared to those with DLD. This difference is statistically significant (*p* = 0.05). More pronounced variations are observed between children with HI and those with TD, as well as between children with DLD and TD. These comparisons reveal significant differences, indicating that deletions are relatively rare in children with TD.

#### 3.1.2 Insertions

Table 5 shows the comparative effects of the three groups, namely DLD, HI, and TD on the phonemic insertion errors.

**Table 5.** Statistical analysis of the effect of the group on the insertions.

| *Effect* | *Estimate* | 95% Confidence Intervals |  |  | *df* | *t* | *p* |
| --- | --- | --- | --- | --- | --- | --- | --- |
|  |  | *SE* | *Lower* | *Upper* |  |  |  |
| (Intercept) | 0.17624 | 0.0651 | 0.0487 | 0.3038 | 2.04 | 2.708 | 0.111 |
| HI - DLD | 0.00253 | 0.0204 | -0.0374 | 0.0424 | 67.59 | 0.125 | 0.901 |
| TD - DLD | -0.04733 | 0.0162 | -0.0792 | -0.0155 | 65.77 | -2.915 | 0.005 |

Notes: DLD = developmental language disorder, HI = hearing impairment, TD = typical development.

The post-hoc Bonferroni analysis showed that children with DLD different significantly compared to those with TD in insertions (*p* = 0.015), primarily because they tended to produce more insertion errors. Similarly, children with HI also differed significantly from those with TD (*p* = 0.052). However, there was no significant difference in the frequency of insertion errors between children with DLD and those with HI.

#### 3.1.3 Substitutions

Table 6 shows the comparative effects of the three groups, namely DLD, HI, and TD on the phonemic insertion errors.

**Table 6.** Statistical analysis of the effect of the group on the phonemic substitution errors.

| *Effect* | *Estimate* | *SE* | 95% Confidence Intervals |  | *df* | *t* | *p* |
| --- | --- | --- | --- | --- | --- | --- | --- |
|  |  |  | *Lower* | *Upper* |  |  |  |
| (Intercept) | 1.6132 | 0.513 | 0.608 | 2.618 | 2.05 | 3.146 | 0.085 |
| HI - DLD | 0.0498 | 0.148 | -0.24 | 0.339 | 69.03 | 0.337 | 0.737 |
| TD - DLD | -0.8347 | 0.119 | -1.067 | -0.602 | 68.61 | -7.033 | < .001 |

Notes: DLD = developmental language disorder, HI = hearing impairment, TD = typical development.

The post-hoc Bonferroni analysis showed that children with DLD differed significantly from those with TD (p < .001) as children with DLD produced more substitution errors that TD children. Children with HI also showed significant differences from those with TD (p < .001). However, there was no significant difference in the incidence of insertion errors between children with DLD and those with HI.

#### 3.1.4 Transpositions

**Error! Reference source not found.** shows the comparative effects of the three groups, namely DLD, HI, and TD on the phonemic transposition errors.

**Table 7.** Statistical analysis of the effect of the group on the phonemic transposition errors.

| Effect | *Estimate* | *SE* | 95% Confidence Intervals |  | *df* | *t* | *p* |
| --- | --- | --- | --- | --- | --- | --- | --- |
|  |  |  | *Lower* | *Upper* |  |  |  |
| (Intercept) | 0.00887 | 0.0035 | 0.00201 | 0.01574 | 2.21 | 2.533 | 0.115 |
| HI - DLD | -0.00111 | 0.00319 | -0.00736 | 0.00514 | 68.65 | -0.349 | 0.728 |
| TD - DLD | -0.00372 | 0.00253 | -0.00868 | 0.00124 | 66.22 | -1.47 | 0.146 |

Notes: DLD = developmental language disorder, HI = hearing impairment, TD = typical development.

We found no errors with regards to transposition errors between the three groups.

### 3.2 Classification 1: TD vs non-TD children

We evaluated three classification models: Decision Trees, Random Forests, and Gradient Boosting. The classification models were able to classify between TD children and non-TD children. The best model, a Random Forests classifier, was able to perform the classification with a relatively high classification accuracy and F1-score, i.e., 93%.

The ROC Curve of the best performing model, namely a RF model, which was 88%. Precision informs the model’s accuracy of positive predictions. The model’s accuracy, precision and F1-score approximated a mean 93% across folds, which indicates good overall performance across both classes. We run a feature importance, which showed that the CPS had substantial contribution.

**Table 2.** The ranking of the first 5 features and their importance.

| Ranking | Feature | Feature Importance |
| --- | --- | --- |
| 1 | CPS | 0.60 |
| 2 | Expressive grammar | 0.17 |
| 3 | Non-Verbal | 0.08 |
| 4 | Non-Word Repetition | 0.07 |
| 5 | Receptive Vocabulary | 0.04 |

Note: Feature importance is total score of 1.

### 3.3 Classification 2: Children with DLD vs. HI

We followed the same model classification approach as in classification 1 and evaluated three classification models: Decision Trees, Random Forests, and Gradient Boosting. The best model in this case was a Decision Tree classifier and classified children with DLD and HI with 91% classification accuracy and F1-score (Table 2) and ROC/AUC was 86% (Figure 2).

**Figure 2.**
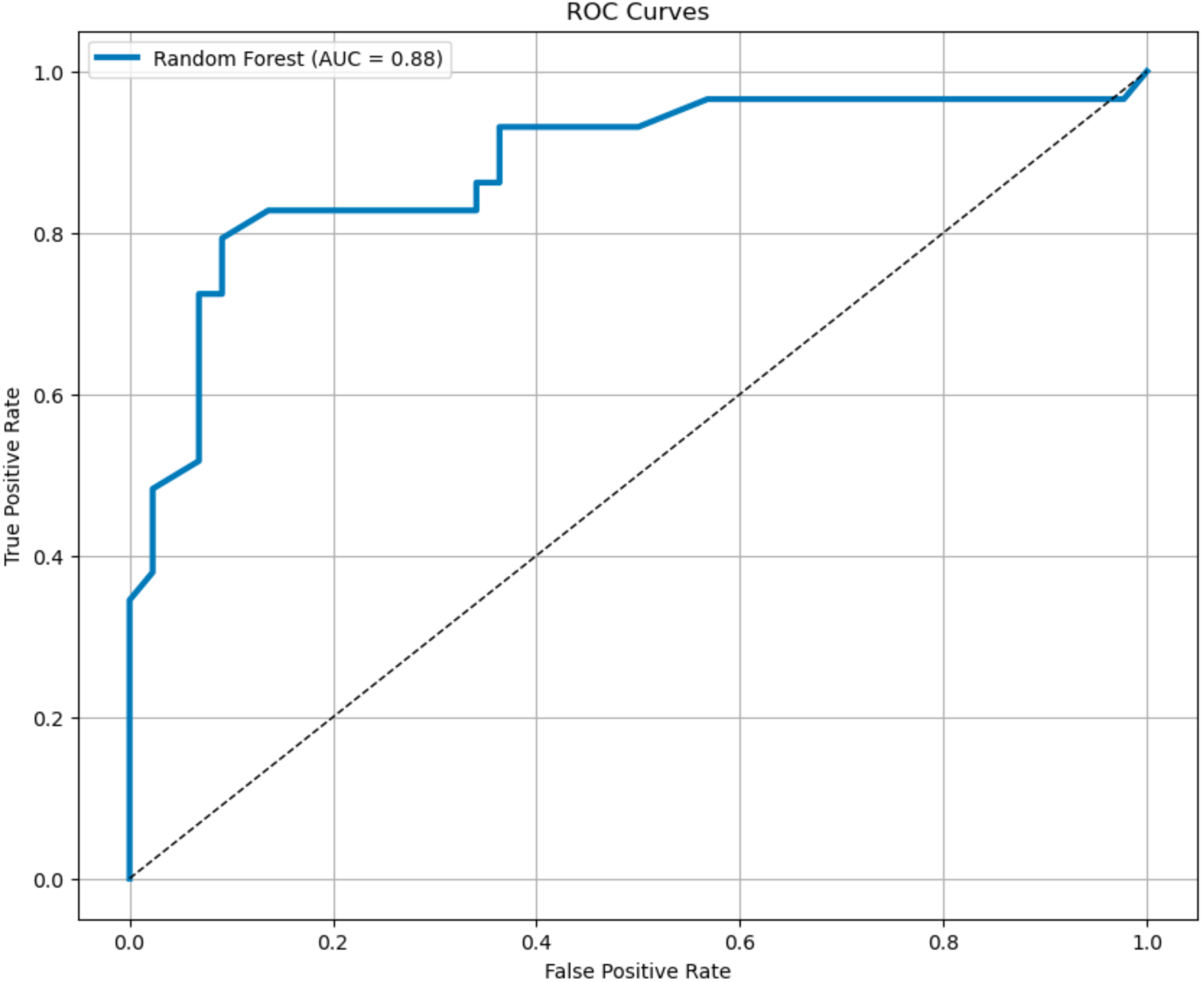
ROC / AUC Curve of the best performing model (AUC = 88%).

**Figure.**
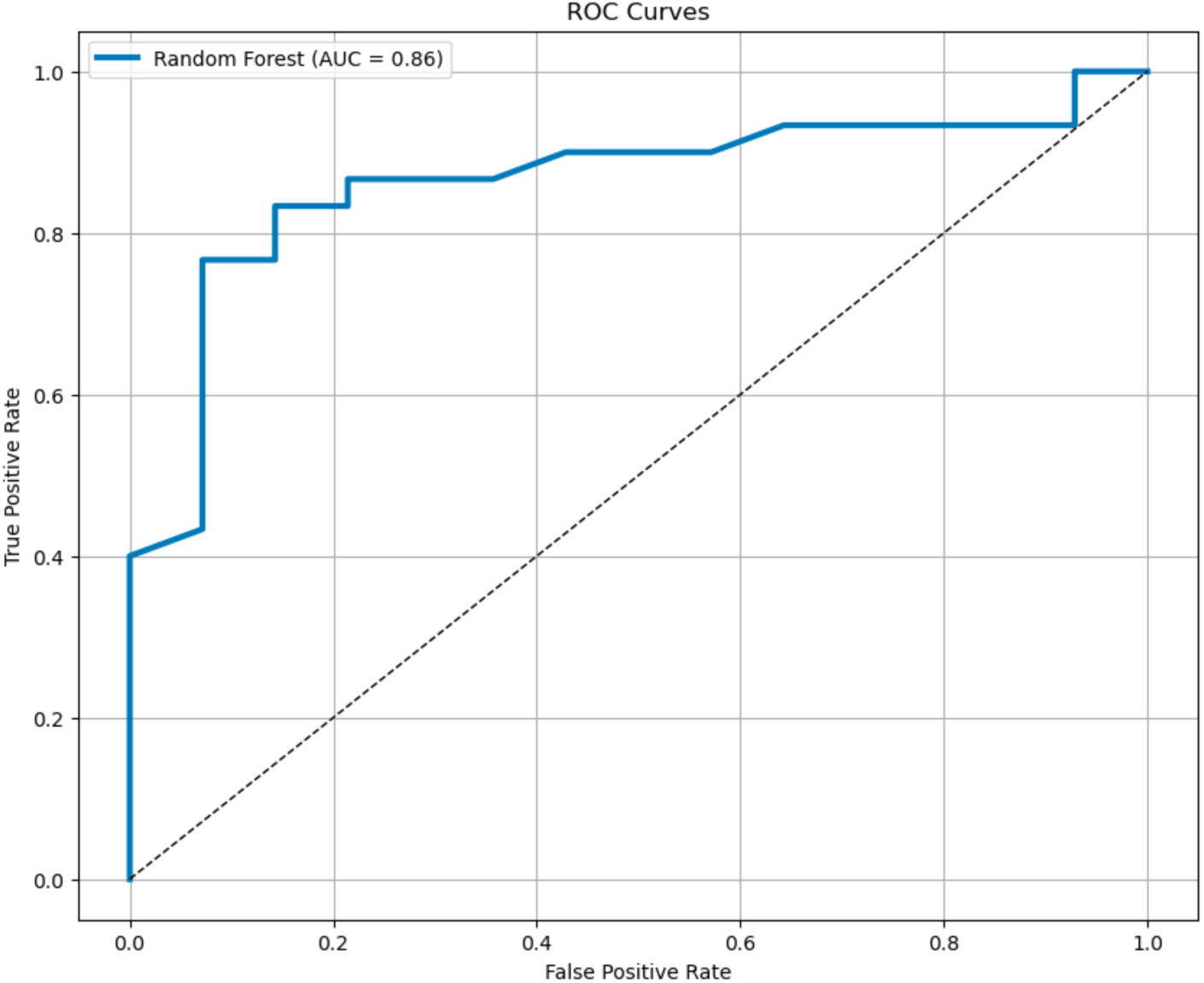
ROC / AUC Curve of the best performing model (AUC = 86%).

Table 8 shows the precision, recall, F1-score, and support for each label, along with the overall accuracy, macro average, and weighted average for the model. The model is highly effective, especially in predicting class 1 with high precision and good recall. Class 0, while having a lower precision, still had excellent recall. The overall accuracy and weighted metrics suggest that the model performs robustly, making it quite effective for tasks where both classes are crucial. The high F1-scores across both classes also indicate a strong balance between precision and recall, crucial for many practical applications. The feature importance is shown in Table 9.

**Table 8.** Classification results of the best performing model, a Random Forest model.

| Label | Precision | Recall | F1-score |
| --- | --- | --- | --- |
| Accuracy |  |  | 0.91 |
| Macro Avg | 0.89 | 0.91 | 0.90 |
| Weighted Avg | 0.92 | 0.91 | 0.91 |

**Table 9.**
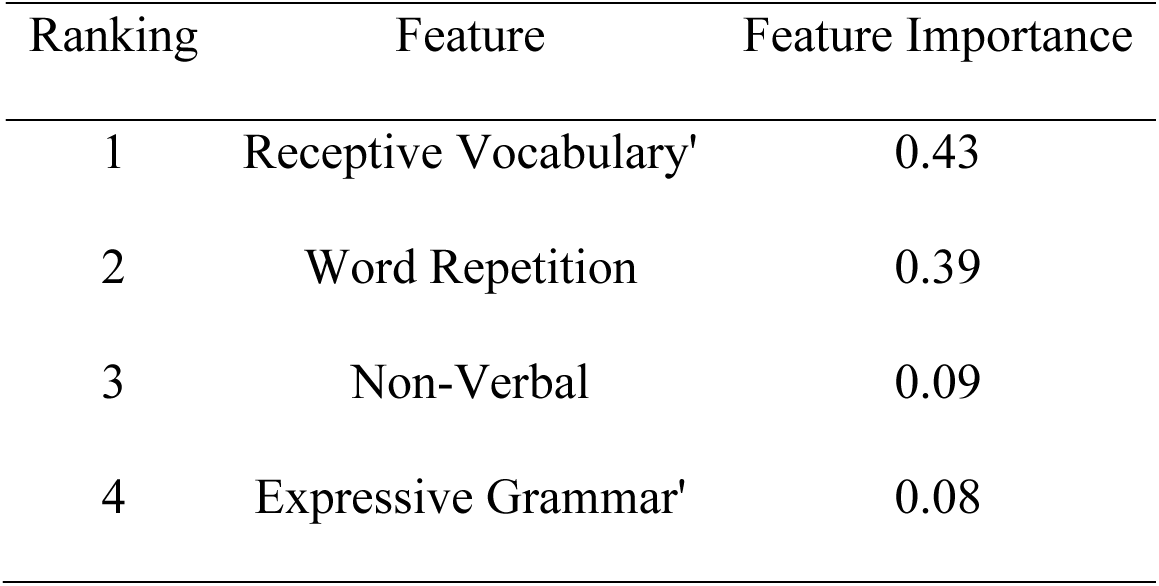
The ranking of the first 5 features and their importance.

| Ranking | Feature | Feature Importance |
| --- | --- | --- |
| 1 | Receptive Vocabulary' | 0.43 |
| 2 | Word Repetition | 0.39 |
| 3 | Non-Verbal | 0.09 |
| 4 | Expressive Grammar' | 0.08 |

The most importance feature was Receptive Vocabulary and Word Repetition. Unlike the previous case, the role of the CPS to distinguish between individuals with DLD and HI was negligeable.

## 4 Discussion

Traditional phonological assessments rely heavily on manual scoring, which is time-consuming, labor-intensive, and prone to human error and variability. This study addresses the challenges of manually scoring phonological errors by proposing an automated system that calculates phonemic errors and introduces a novel approach by leveraging computational methods to standardize and streamline this process (Heeringa et al., 2009; Hixon et al., 2011; Riches et al., 2011; Smith et al., 2019). The new algorithm calculates a Compositive Phonological Score (CPS) from the Normalized Damerau–Levenshtein Distance for phonemic error calculation and, at the same time, introduces specific scores for individual errors like deletions, insertions, substitutions, and transpositions (Themistocleous, 2024; Themistocleous et al., 2020). The proposed automated system ensures consistency across assessments by applying the same criteria and calculations to each case, eliminating the subjectivity inherent in manual scoring. This consistency is crucial for reliable diagnosis and tracking of progress over time. Also, the automated scoring enhances precision in phonemic error analysis, as the system can measure phonemic differences between a child’s production and the target pronunciation, providing detailed insights that are difficult to achieve manually.

Specifically, we have applied this algorithm to assess phonological performance in Swedish-speaking children with DLD and HI, compared to TD children, and provided a detailed analysis of phonological errors among Swedish-speaking children with DLD, HI, and TD children, demonstrating distinct patterns of phonological errors across these groups (Sundström et al., 2018a; Sundstrom et al., 2019). This study had two primary objectives. First, we aimed to quantify phonological production differences in Swedish-speaking children with DLD and HI compared to TD children. The study demonstrated that children with DLD and HI exhibited significantly higher CPS than TD children, indicating more frequent phonological errors. DLD and HI groups showed a CPS of around 0.27-0.28, while TD children had a lower CPS of 0.11. The findings suggest that phonological errors are considerably more common in children with language impairments than in typically developing peers (Andreou et al., 2024).

However, the CPS did not significantly differ between the DLD and HI groups, suggesting similar levels of overall phonological difficulty in both groups. The lack of significant difference between DLD and HI groups underscores the complexity of diagnosing and treating language disorders that manifest similar phonological difficulties (Law et al., 1996). It highlights the need for comprehensive and multifaceted approaches to diagnosis and intervention, the importance of understanding these conditions’ shared and unique aspects, and the potential challenges in clinical practice. Also, the phonological difficulties in DLD and HI groups underscore that interventions targeting phonological errors might need to be similar for both groups. However, since the underlying causes of these errors differ (DLD being a language disorder and HI related to hearing loss), interventions should still be tailored to address each group’s specific needs and underlying issues. For children with HI, interventions can focus more on improving auditory processing and maximizing residual hearing through assistive technologies (e.g., hearing aids and cochlear implants). In contrast, for children with DLD, therapy might emphasize enhancing language processing and production skills (Hoover, 2019). The similarity in CPS underscores the need for interventions to go beyond phonological errors and address the broader context of each disorder.

Concerning the specific types of Phonological Errors, substitutions were the most common error across all groups, with DLD and HI children showing significantly higher rates than TD children. Thus, replacing one phoneme with another is a prevalent issue in DLD and HI. Deletions were more frequent in the HI group than in the DLD and TD groups, with the HI group showing a trend towards more deletion errors than the DLD group. Both DLD and HI groups had significantly more deletions compared to TD children. Insertions were relatively rare across all groups, but both DLD and HI children exhibited higher rates than TD children. The differences between DLD and TD were significant, highlighting that children with language disorders may add extra phonemes do not present in the target word. Transpositions were the most minor common type of error and did not show significant differences between the groups, indicating that swapping adjacent phonemes is not a prominent issue in these populations. These findings indicate that despite different etiologies, children with DLD and HI experience comparable difficulty levels in phonological production, which suggests that phonological errors in these conditions may stem from broader, shared difficulties in language processing, whether due to impaired auditory input (in HI) or impaired linguistic processing (in DLD). Understanding these shared challenges can inform the development of more effective cross-cutting intervention strategies.

Second, the study intended to develop a classification model to differentiate between children with DLD, HI, and aged matched typically developing children based on their phonological production patterns. We hypothesized that Swedish-speaking children with DLD and HI will exhibit distinct phonological production profiles compared to TD children and differ from each other. The hypothesis was confirmed, and the Classification of TD vs. Non-TD Children resulted in a high classification accuracy. Specifically, the Random Forest classifier was the most effective, achieving high classification accuracy (93%) and a strong F1 score. The CPS emerged as the most crucial feature in distinguishing TD children from those with language impairments (DLD and HI), underscoring the utility of the CPS metric in clinical diagnostics. The classification results of DLD vs. HI Children showed that the Decision Tree classifier had the best performance, with an accuracy of 91% and a high F1 score.

Interestingly, in this classification, the CPS was not a significant feature. Instead, Receptive Vocabulary and Word Repetition were the most important predictors, which suggests that while CPS is crucial for identifying general language impairment, other linguistic measures are more critical in differentiating between specific disorders like DLD and HI.

The study offers a more objective and consistent method for assessing phonological performance by quantifying phonological errors with the CPS. The method can reduce reliance on labor-intensive manual scoring and minimize the risk of human error, making diagnostics more precise and accessible. The study provides valuable insights into the phonological profiles of children with DLD and HI. The several types of phonological errors highlight the specific challenges faced by these groups, such as a higher prevalence of substitutions and deletions. These findings can inform targeted intervention strategies that address the needs of children with these language impairments. The study’s application of machine learning models to classify children based on their phonological errors is another significant contribution as it demonstrates the potential that the combination of metrics employed in this study can corroborate existing screening approaches for DLD and HI (see Bao et al. (2024) for a recent review). The ability to accurately distinguish between TD, DLD, and HI children using phonological metrics and other linguistic measures demonstrates the potential of AI in enhancing early diagnosis and intervention. This approach can be particularly beneficial in resource-limited settings where access to specialized clinicians is scarce.

### 4.1 Limitations and Future Research

While this study contributes significantly to phonological assessment and language disorder diagnosis, several limitations should be noted. First, the study involved a relatively small sample size of 72 children, which may limit the generalizability of the findings. Although the study included three distinct groups (DLD, HI, and TD), the small number of participants in each group may not fully capture the variability in the broader population of children with these conditions. The study is also limited to Swedish-speaking children, specifically those speaking the Central Swedish dialect. This focus restricts the applicability of the findings to other languages or dialects, where phonological rules and error patterns may differ. Finally, the proposed method capture only phonemic level differences, thus, domains such as lexical and post-lexical prosody, which might be affect in children with HI and TD require a separate scoring algorithm (Sundstrom et al., 2019; Themistocleous, 2016). Despite these limitations, the study offers valuable insights and tools for assessing phonological errors in children with language disorders.

Future research should aim to address these limitations by including larger and more diverse populations, exploring additional linguistic factors, and refining the machine learning models and error analysis methods to enhance the robustness and applicability of the findings. By establishing measurable benchmarks for phonological errors, the study contributes to the early diagnosis and treatment of language disorders. Early identification of children with DLD or HI can lead to timely interventions, potentially mitigating the long-term impacts of these disorders on language development and academic achievement (Davidson et al., 2019). The study’s implementation of the phonological scoring algorithm in Open Brain AI (http://openbrainai.com), with multilingual support, underscores its broad applicability (Themistocleous, 2024). This makes the tool feely accessible to a wide range of clinicians and researchers, facilitating its integration into various linguistic and cultural contexts. This generalizability enhances the tool’s utility in global clinical practice and research.

### 4.2 Conclusion

The study represents a significant advancement in the automated assessment of phonological errors, offering a reliable, scalable tool for diagnosing and understanding language disorders. Its integration of machine learning further enhances its diagnostic power, making it a valuable resource for clinicians and researchers aiming to improve outcomes for children with language impairments.

## Data Availability Statement

The data are available upon request to the first author (SS). The Phonological Scoring Application, developed by the second author, is freely accessible at Open Brain AI (http://openbrainai.com).

